# A combined oro-nasopharyngeal swab is more sensitive than mouthwash in detecting SARS-CoV-2 by a high-throughput PCR assay

**DOI:** 10.1101/2020.09.25.20201541

**Authors:** Wiebke Michel, Jacqueline Färber, Milica Dilas, Ina Tammer, Jannik Baar, Achim J. Kaasch

**Affiliations:** Otto-von-Guericke-university Magdeburg, Medical faculty, Institute of Medical Microbiology and Hospital Hygiene

## Abstract

**Objectives:** The optimal diagnostic specimen to detect SARS-CoV-2 by PCR in the upper respiratory tract is unclear. Mouthwash fluid has been reported as an alternative to nasopharyngeal and oropharyngeal swabs. We compared mouthwash fluid with a combined oro-nasopharyngeal swab regarding test performance.

**Methods:** We tested asymptomatic persons with a previous diagnosis of COVID-19 and their household contacts. First, a mouthwash (gargling for at least 5 sec) with sterile water was performed. Then, with a single flocked swab the back of the throat and subsequently the nasopharynx were sampled. Samples were inactivated and analysed on a Roche cobas 6800® system with the Roche SARS-CoV-2 test.

**Results:** Of 76 persons, 39 (51%) tested positive for SARS-CoV-2 by oro-nasopharyngeal swab. Mouthwash detected 13 (17%) of these infections but did not detect any additional infection. Samples that were positive in both tests, had lower cycle threshold (Ct)-values for oro-nasopharyngeal samples, indicating a higher virus concentration, compared to samples only positive in oro-nasopharyngeal swabs.

**Conclusions:** Mouthwash is not as sensitive as combined oro-nasopharyngeal swab in detecting upper respiratory tract infection.

## Introduction

In December 2019, a new lung disease called Coronavirus Disease 2019 (COVID-19) first appeared in Wuhan, China, and subsequently spread globally [1]. The causative agent is the severe acute respiratory syndrome coronavirus 2 (SARS-CoV-2). SARS-CoV-2 is an enveloped, single-stranded positive-sense RNA virus. Together with SARS-1 and MERS coronavirus it is classified in the *Orthocoronaviridae* subfamily, genus *Betacoronavirus* [2].

SARS-CoV-2 is efficiently transmitted from person to person by respiratory droplets [3, 4]. Rapid and accurate detection of the virus is essential to contain outbreaks. The most suitable diagnostic specimen is still unclear, as the virus is detectable in different respiratory specimens, urine, and stool [5, 6]. One recommended diagnostic specimen for SARS-CoV-2 detection is the nasopharyngeal swab [7], but combined naso-oropharyngeal swabs can increase the sensivity of SARS-CoV-2 detection [4]. A meta-analysis of different SARS-CoV-2 studies showed the highest detection rates in sputum, followed by nasopharyngeal and then oropharyngeal swab samples [8]. In severe cases of COVID-19 or at later stages in the disease, SARS-CoV-2 can be detected in samples from the lower respiratory tract, such as sputum or bronchial aspirate [9].

Expected shortages of swabs led us to assess alternative diagnostic specimens. In this study, we compared test performance when using mouthwash or a combined oro-nasopharyngeal swab.

## Methods

Residents (age >6 years) from a refugee facility with a previous diagnosis of COVID-19 and their household contacts were prospectively tested for SARS-CoV-2 with mouthwash and a combined oro-nasopharyngeal swab in controlled conditions on two occasions in May 2020 during an outbreak in the facility. Symptoms of COVID-19 were recorded using a standardized questionnaire. Samples were taken by previously instructed medical personnel. A single flocked swab (eSwab™ Copan) was used to sample the back of the throat and subsequently the deep nasopharynx. For the mouthwash, residents were instructed to gargle the mouth with 10 ml sterile water for at least 5 seconds. Samples were transported at room temperature and stored overnight at 4°C. All samples were mixed 1:1 with ATL buffer and analysed with the cobas® SARS-CoV-2 assay on the Roche cobas 6800 system according to the manufacturer’s instructions. Detection of the *E*-(envelope)-gene and *Orf1/a* (open reading frame 1) or only *E*-gene or only *Orf1/a* were interpreted as confirmation of SARS-CoV-2 infection. Cycle threshold (Ct)-values above 40 were considered as negative.

The study was performed according to the principles of the Declaration of Helsinki. Approval was obtained from the ethics committee of the Medical Faculty of the Otto-von-Guericke University Magdeburg. Written informed consent was obtained by all participants or their guardians.

## Results

Overall, 64 asymptomatic persons with a previous diagnosis of COVID-19 and their household contacts were tested on two occasions. Age ranged from 7 to 59 years, with an average age of 29 years. At the time of testing, no person showed symptoms of COVID-19. Fifteen persons recollected symptoms compatible with COVID-19 in the past three weeks: cough (6 persons), headache (4 persons), rhinitis (4 persons), loss of taste (2 persons), fatigue (2 persons), fever, sneezing, aching limbs, diarrhoea, and mild shortness of breath (one person each). No person was hospitalised.

In 39 of the 76 participants (51%), PCR of the combined oro-nasopharyngeal swab confirmed SARS-CoV2 infection. In contrast, SARS-CoV-2 was detected in 13 mouthwashes (17 %) of which all were positive by the oro-nasopharyngeal swab (table 1). When considering the oro-nasopharyngeal swab as gold standard, the sensitivity of mouthwash was 33%.

**Table 1:**
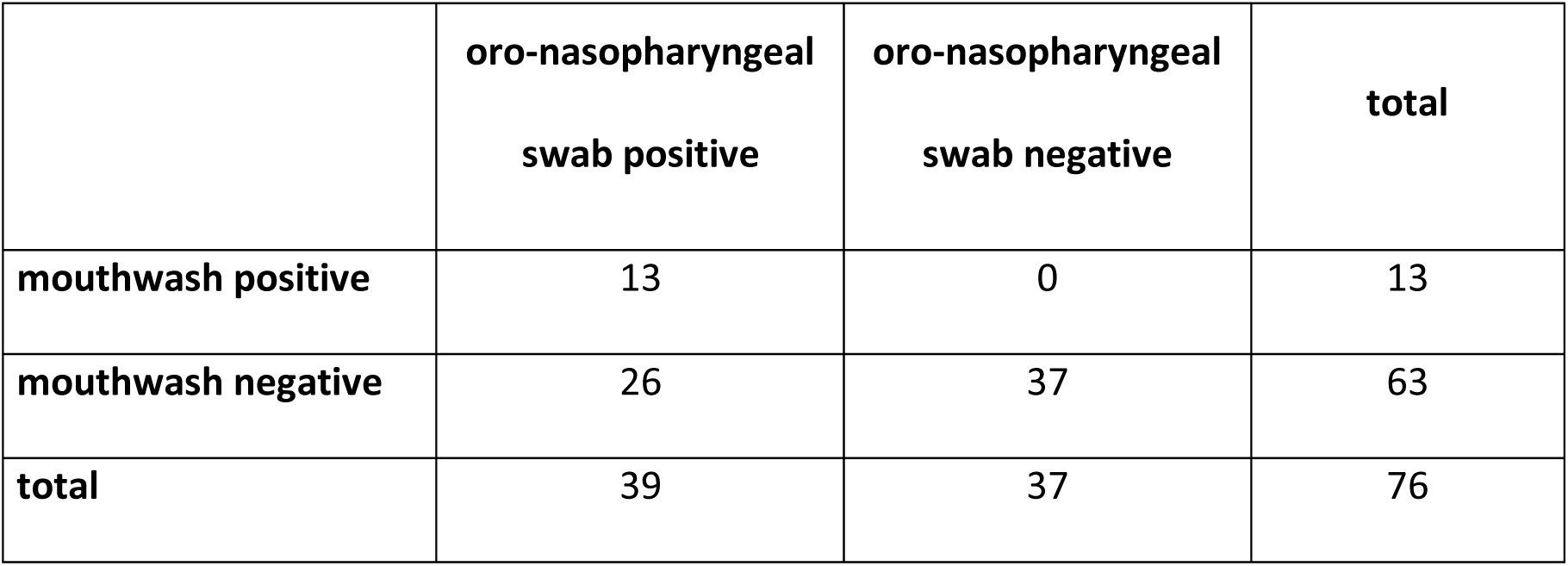
Comparison of mouthwash and combined oro-nasopharyngeal swab in detecting SARS-CoV-2. The sensitivity of mouthwash is 33%, the specificity 100% when using the combined oro-nasopharyngeal swab as gold standard (McNemar test *p*-value <0.001).

The cycle threshold (Ct)-value is a measure for the abundance of the transcript in the sample and correlates with viral load. When comparing Ct-values of oro-nasopharyngeal swabs, specimens that were positive by mouthwash had lower Ct-values than specimens negative by mouthwash, indicating a lower viral load in mouthwash (Figure 1a). Samples that were positive by both methods showed higher Ct-values in the mouthwash (Figure 1b). All results are consistent with a lower sensitivity of detection in mouthwash.

**Figure 1:**
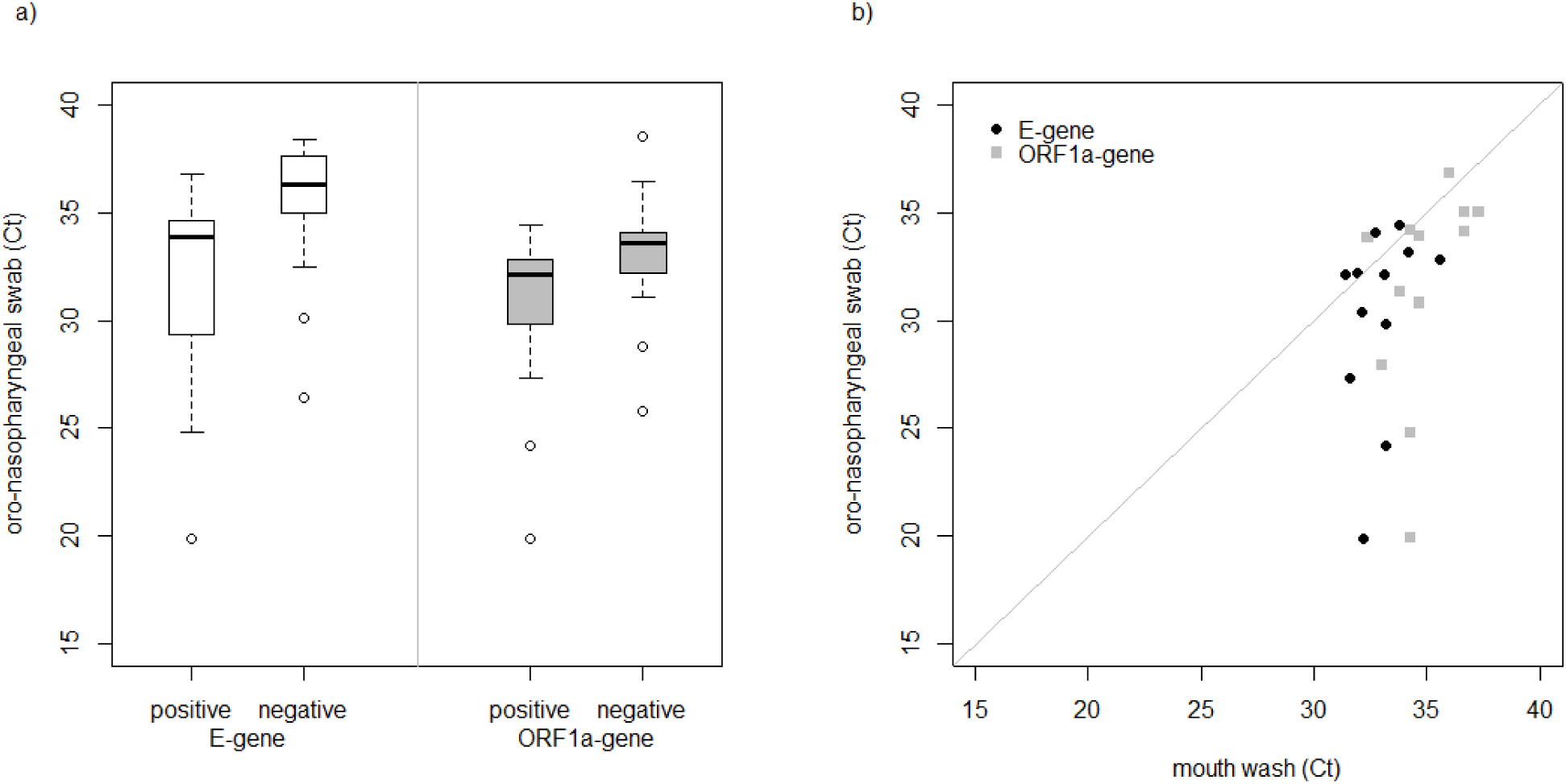
The combined oro-nasopharyngeal swab has a higher sensitivity than mouthwash. a) Cycle threshold (CT) values for *Orf1/a*- and *E*-gene for oro-nasopharyngeal swabs in samples positive and negative in mouthwash. A lower CT-value indicates a higher viral load. (Mann-Whitney U-Test *E*-gene: p-value<0.001, *Orf1/a*-gene: p-value=0.036) b) Ct-values for samples positive in both specimen types. Ct-values for the mouthwash were higher than for the combined oro-nasopharyngeal swabs, indicating a lower viral load in mouthwash. Only 12 paired samples were shown, since one sample was positive in the *E*-gene and another in the *Orf1/a*-gene, only (Wilcoxon signed rank test *E*-gene: p-value=0.007, *Orf1/a*-gene: p-value=0.037).

## Discussion

The shortage of swabs that are suitable for PCR diagnostics led us to explore the utility of mouthwash in a controlled study. We found a very low sensitivity of mouthwash (33%), when using oro-nasopharyngeal swabs as comparator. We speculate that this striking difference in sensitivity is partly due to the dilution of the mouthwash sample. Thus, mouthwash is not suitable for the reliable detection of SARS-CoV-2 infection.

The only published study reported that the rate of positivity for SARS-CoV-2 was higher in self-collected throat washings with sterile normal saline than in nasopharyngeal swabs [10]. However, the small sample size of eleven patients does not allow firm conclusions.

Our study has several strengths: We conducted the study in a controlled setting with specifically trained personnel. This allows for a more rigorously sampling than in an observational study conducted in the clinical setting. As gold standard, we chose combined oro-nasopharyngeal swabs. A systematic review that assessed the positivity rate of different specimens found that nasopharyngeal swabs had a slightly higher positivity rate than oropharyngeal swabs, with larger differences when sampling was performed more than 14 days after symptom onset [8].

Our study population were asymptomatic persons, with a median time after diagnosis of 14 days, and their household contact. Since the viral load decreases over time, this population is expected to have a low viral load and thus high Ct-values. Indeed, 34 of 38 (89%) samples had Ct-values above 30 for the *E-*gene, a value currently discussed as a cut-off for infection. Thus, this study was designed to rigorously assess differences in sensitivity.

Our study has also limitations. Mouthwash with gargling was performed as a self-administered procedure and we observed some variation in adherence to the protocol regarding the duration and intensity of gargling, which may have influenced the results. Furthermore, we did not compare different RNA extraction methods, which may show a better performance with mouthwash specimens.

There is a high likelihood of aerosol formation during gargling. Thus, mouthwash should be performed alone in a well-ventilated area. This may limit its use in patients to minimize exposure of health-care personel.In conclusion, SARS-CoV2 detection with mouthwash showed a low sensitivity compared to oro-nasopharyngeal swabs. Thus, we do not recommend mouthwash performing combined oro-nasopharyngeal swabs, especially in patients with no or mild symptoms.

## Transparency declaration

JF reports personal fees from Biomérieux and the Medical Association of Saxony-Anhalt, outside the submitted work; WM, MD, IT, JB and AJK report no conflicts of interest. There was no specific funding for this study.

## Data Availability

Data available on request.

## Contribution

AJK and IT designed the study; DM, JB, IT and AJK conducted the investigation; MW, BJ, and AJK edited, reviewed and interpreted the data; WM and AJK wrote the original draft and all authors approved the manuscript.

## Acknowledgement

We thank the technical personnel of the Institute of Medical Microbiology and Hospital Hygiene and the Institute of Transfusion Medicine, Medical Faculty, Otto-von-Guericke University, for technical support.

